# Multi-modality detection of SARS-CoV-2 in faecal donor samples for transplantation and in asymptomatic emergency surgical admissions

**DOI:** 10.1101/2021.02.02.21250934

**Authors:** Susan E Manzoor, Shafquat Zaman, Celina Whalley, David Inglis, Andrew Bosworth, Michael Kidd, Sahida Shabir, Nabil Quraishi, Christopher A Green, Tariq Iqbal, Andrew D Beggs

**Author notes:** Correspondence to: Professor Andrew Beggs, Institute of Cancer & Genomic Sciences, University of Birmingham, Vincent Drive, Birmingham, B15 2TT, United Kingdom. Authors contributed equally to the manuscript (Tariq Iqbal, Andrew D Beggs). Role of funders: This report presents independent research and the views expressed in this publication are those of the authors and not necessarily those of the NHS, or the Department of Health and Social Care. Competing interests: ADB – has received travel funding to the Oxford Nanopore Community Meeting 2019 from Oxford Nanopore. The rest of the authors declare no competing interests. Authorship statement: Study conception: ADB, TI, CAG Sample collection: SM, SZ, SS, DI, TI Molecular analysis: SM, SZ, CW, AB, MK Data analysis: all authors Paper writing: all authors Guarantor: ADB.

## Abstract

**Introduction:** Faecal transplantation is an evidence based treatment for *Clostridiodes difficile*. Patients infected with SARS-CoV-2 have been shown to shed the virus in stool for up to 33 days, well beyond the average clearance time for upper respiratory tract shedding. We carried out an analytical and clinical validation of reverse-transcriptase quantitative (RT-qPCR) as well as LAMP, LamPORE and droplet digital PCR in the detection of SARS-CoV-2 RNA in stool from donated samples for FMT, spiked samples and asymptomatic inpatients in an acute surgical unit.

**Methods:** Killed SARS-CoV-2 viral lysate and extracted RNA was spiked into donor stool & FMT and a linear dilution series from 10^−1^ to 10^−5^ and tested via RT-qPCR, LAMP, LamPORE and ddPCR against SARS-CoV-2. Patients admitted to the critical care unit with symptomatic SARS-CoV-2 and sequential asymptomatic patients from acute presentation to an acute surgical unit were also tested.

**Results:** In a linear dilution series, detection of the lowest dilution series was found to be 8 copies per microlitre of sample. Spiked lysate samples down to 10^−2^ dilution were detected in FMT samples using RTQPCR, LamPORE and ddPCR and down to 10^−1^ with LAMP. In symptomatic patients 5/12 had detectable SARS-CoV-2 in stool via RT-qPCR and 6/12 via LamPORE, and in 1/97 asymptomatic patients via RT-qPCR.

**Conclusions:** RT-qPCR can be detected in FMT donor samples using RT-qPCR, LamPORE and ddPCR to low levels using validated pathways. As previously demonstrated, nearly half of symptomatic and less than one percent of asymptomatic patients had detectable SARS-CoV-2 in stool.

## INTRODUCTION

Severe acute respiratory syndrome coronavirus 2 (SARS-CoV-2) infection, which causes coronavirus disease 2019 (COVID-19), first emerged in Wuhan, China in late 2019 (1) and is responsible for nearly 1.5 million deaths worldwide. SARS-CoV-2 is primarily transmitted via respiratory droplets and direct contact routes between asymptomatic and symptomatic individuals. Although faeco-oral transmission has not been documented with this virus (2), it is one of the main forms of transmission with other similar single-stranded RNA viruses, such as norovirus (3). Multiple lines of evidence suggest that the gastrointestinal tract may form a reservoir for SARS-CoV-2 with the potential for infection and transmission(4). The virus enters the host by binding its S1 “spike” glycoprotein to angiotensin-converting enzyme 2 (ACE2) in epithelial tissues which is avidly expressed in the ileum (5). Furthermore, gastro-intestinal symptoms such as diarrhoea are not uncommon in patients infected with SARS-CoV-2 affecting up to 40% patients admitted to hospital. Several studies have identified that over 40% of patients with detectable SARS-CoV-2 RNA from nasopharyngeal swabs, will also have detectable viral RNA in faecal samples(6). Moreover, faecal shedding of viral RNA has been shown to persist for up to 33 days after clearance from respiratory samples(6). It is not known for certain whether SARS-CoV-2 present in faeces represents live and transmissible virus (7), although early evidence suggests that this is possible in some and there remains uncertainty regarding the role of the gut in COVID-19 pathogenesis, potential for faeco-oral transmission of the virus and future outbreaks of infection in institutions such as hospitals and care homes.

Faecal microbiota transplantation (FMT) involves the transplantation of processed faecal samples (8) from healthy donors to individuals with disease associated with imbalance in the gut microbiome. In recent years FMT has transformed the treatment of patients with *Clostridiodes difficile* infection (9), especially in those with recurrent or refractory disease. Although FMT donors and their stool donations undergo screening to good manufacturing procedure (GMP) standards for pathogens of potential significance, such as multi-drug resistant *Enterobacteriaceae*, some risks of this remain (10). During the current pandemic many stool banks have need to stop providing FMT due to the potential risk of transmission of SARS-CoV-2 from asymptomatic donors and, earlier this year the Federal Drug Administration mandated that only stool donated prior to 1^st^ December 2019 could be used for FMT. A recent international consensus paper recommenced direct stool testing for the presence of SARS-CoV-2 would be needed for safe FMT supply in the COVID-19 era 2020 (11, 12).

In this paper we present the first the results of testing FMT donor stool for the presence of SARS-CoV-2 virus using an internally developed assay based on an existing CE-IVD marked product as well as various novel diagnostics that have been developed in response to the pandemic. We also report using this to test asymptomatic acute patients admitted to hospital with COVID-19 as part of the ‘second spike’ of the pandemic in 2020 in order to understand the prevalence of faecal SARS-CoV-2 detection in this cohort.

## METHODS

### Patient samples

Stool samples were collected under an existing gut microbial profiling study with ethical approval from Yorkshire & The Humber - Bradford Leeds Research Ethics Committee (16/YH/0100). Samples were obtained from 12 symptomatic COVID-19 patients admitted to hospital who tested positive for SARS-CoV-2 RNA with PCR testing on naso-pharyngeal swabs and a further 97 asymptomatic patients presenting to the Surgical Assessment Unit of Sandwell and West Birmingham NHS Trust from September-November 2020 (a time of increasing prevalence).

### Stool and FMT samples

Samples used for spiking experiments were taken from pre-existing stocks at the University of Birmingham Microbiome Treatment Centre FMT bank. These were from a batch collected and stored in 2017, prior to the emergence of the SARS-CoV-2 virus. These samples were collected in readiness for use in a trial of FMT in inflammatory bowel disease (STOP-COLITIS 2015-005753-12). For this study, FMT was manufactured over a 10-day donation period and retention and stool study samples were collected and stored in accordance with an MHRA approved GMP process REC and ethical approval (17/EM/0274).

### Viral spiking experiments

Quantitative PCR was used to analyse both from COVID-19 patients and pre-COVID FMT following “spiking”, with varying concentrations of SARS-CoV-2 inactivated lysate (cell culture in Qiagen AL buffer). Purified RNA extracted from an aliquot of the lysate was also analysed using quantitative PCR as an additional positive control to determine whether RNA could be recovered and detected, and to determine the limit of the assay. A negative control of nuclease-free water was incorporated into the assay.

Live SARS-CoV-2 England/2/2020(VE6-T) virus was isolated from infected VE6-T cells, then inactivated in Qiagen lysis buffer AL and frozen at -80°C. Viral RNA was purified from 500μL of cell lysate using a QIAmp Qiagen kit, then diluted serially to 10^−5^ in nuclease free water. Viral cell lysate was also diluted to 10^−5^ serially ten-fold in nuclease free water.

This dilution series of both Lysate and RNA were then used to spike aliquots of faecal stool and FMT samples in duplicate. Extraction control samples consisting of 10-fold dilutions of cell lysate and RNA were also prepared.

### RNA extraction

RNA was extracted from 0.2-0.25g of stool. All stool and FMT ‘spiked’ samples, along with the extraction control samples were lysed using bead beating PM1 buffer (containing guanidinium thiocyanate, Qiagen) to inactivate the virus prior to extraction under containment level 2+ (CL2+) conditions. A RNeasy PowerMicrobiome kit (Qiagen, Hilden) was then used for viral RNA extraction and purification of total RNA from stool with on-column DNase treatment. A subset of samples underwent quality assessment using an Agilent Tapestation 2200 with RNA Broad Range (BR) detection kit in order to understand the effects of RNA extraction upon stool samples.

### Real-Time PCR (RT-PCR)

Two real-time PCR detection kits for the detection of SARS-CoV-2 were used for the detection of virus in FMT and faecal samples and the extraction control samples: VIASURE SARS-CoV-2 Real Time PCR Detection kit (CerTest Biotec S.L.)(13) and the WHO *E* Gene Assay Test (14) in accordance with manufacturer instructions.

Viral RNA extracted from the spiked samples and extraction control samples underwent reverse-transcriptase quantitative polymerase chain reaction (RT-qPCR) using primers against the *orf1ab* polyprotein, nucleoplasmid structural protein (N)and the envelope membrane protein (E)) which have been previously validated for the detection of SARS-CoV-2 (15).

The level of fluorescence produced was analysed using ThermoFisher Connect (TM) software, and a sample was considered positive if the Ct value obtained was less than 38 in any of the gene targets (*ORF1ab, N* or *E*). Positive and negative controls were used in all reactions as well as an RNA extraction control, consisting of spiked viral lysate in buffer AVL.

### Droplet Digital PCR (ddPCR)

Droplet digital real-time PCR assays using target RNA, primers (450nM), fluorescent-labelled probes (200nM) and Bio-Rad ddPCR supermix were prepared. For use in this protocol, the *E* gene primers and 6-FAM labelled probe (used in the WHO *E* gene assay test (14)) were used with the One step RT-ddPCR Advanced Kit (BioRad Laboratories). Samples were fractioned into 20,000 nanolitre sized water-in-oil droplets using QX200 Droplet generator (BioRad Laboratories). The droplets were transferred in a 96-well plate to a thermal cycler where PCR amplification of the template occurs within each droplet. Following PCR each droplet was analysed individually on the QX200 Droplet Reader (BioRad Laboratories) and the fraction of PCR-positive and PCR-negative droplets in the original sample were counted using QuantaSoft software. The data were analysed using Poisson distribution statistics to determine the concentration of target DNA in the original sample in absolute copies/ml.

### LamPORE SARS-CoV-2 detection

The LamPORE SARS-CoV-2 assay utilises a combination of reverse transcriptase loop-mediated isothermal amplification (RT-LAMP) and Nanopore sequencing technology (Oxford Nanopore Technologies, Oxford), as described previously (16, 17). For the SARS-CoV-2 the target regions *N2, E1* and *ORF1a* genes each span approximately 180bp of the viral genome. Briefly, the viral RNA genome was reverse transcribed into cDNA which was then amplified using strand-displacement polymerase to generate concatenated copies of the original target region. The DNA sequences produced (a 2kb concatamer with sequence for 180bp target region) were then aligned against the SARS-CoV-2 genome using a custom algorithm.

### Loop Mediated Isothermal Amplification (LAMP) detection

In order to understand the utility of a rapid assay to detect SARS-CoV-2 in stool, extracted faecal RNA underwent testing using the Optigene RNA-LAMP kit targeted against *ORF1ab* under manufacturer’s instructions for use.

## RESULTS

### Effect of preparation method

To evaluate the analytical sensitivity of the assay and determine the analytical limit of detection, virus derived from *in vitro* cell culture was prepared in two different ways. Lysate was created, by adding equal volumes of cell culture supernatant containing live virus to a Buffer AL (Qiagen), a lysis buffer preparation containing guanidinium thiocyanate designed to both inactivate the virus rendering it safe to handle, and to stabilise RNA to protect it from RNAse mediated degradation.

It is recognised that re-extraction of purified RNA reduces the yield of RNA recovered, and unprotected RNA spiked into a matrix such as stool or FMT preparation is likely to degrade rapidly. To prevent this lysate preparation was also spiked into the stool and FMT preparations. The lysate is less vulnerable as viral RNA is associated with proteins that serve to shield the virus and the addition of guanidinium thiocyanate even at low quantities serves to reduce the activity of RNAse degradative enzymes.

Furthermore, the preparation method for extracting the RNA from the stool and FMT specimens requires mechanical homogenisation, which serves to further fragment and degrade RNA, even in samples such as that introduced in the lysate. When fragment analysis was carried out, we found RNA integrity number (RIN) of <2 in all samples, suggesting RNA processed in this way is highly degraded.

### Analytical sensitivity and specificity

As anticipated spiked RNA into faecal samples yielded poor results (Tables 2 & 3), while the lysate was detectable at 3/5 dilutions. RNA extracted from the FMT and stool where lysate was used as the ‘spike in’, was then tested using RT-ddPCR to determine the copy number recovered in this experiment. The lowest copy number recoverable from this dilution series was 8 copies of virus genome per reaction (or 0.4 copies per microliter of recovered RNA), equating to a limit of detection for the workflow (extraction and ViaSure RT-qPCR) of 8 copies per microliter of recovered RNA. The approximate limit of detection based on this data can be expressed as copies per gram of faecal preparation and is 204 copies of virus in 1g of faecal preparation using the following calculation:

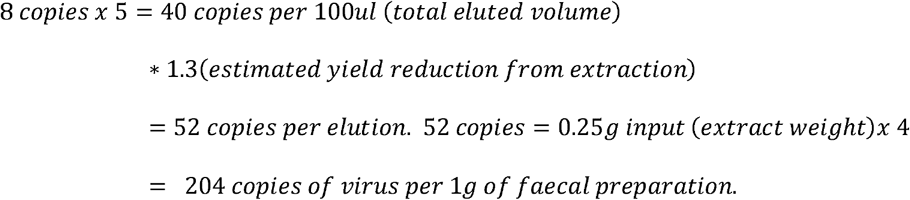

**Table 1.** Clinical data from 12 COVID-19 positive patients

| **Pt. Identifier** | **Gender** | **Presenting Sx** | **GI Sx** | **ITU admission required** | **Treatment Administered** | **Pt. Outcome** |
| --- | --- | --- | --- | --- | --- | --- |
| P01 | F | COUGH / PYREXIA | YES | No | ANTIBIOTICS | DISCHARGED |
| P02 | F | NO DATA | ? | No | NO DATA | DISCHARGED |
| P03 | M | NONE | NO | NO | NONE | DISCHARGED |
| P04 | F | Cough / SOB / Myalgia | YES | No | OXYGEN / ANTIBIOTICS / STEROIDS | DISCHARGED |
| P05 | M | COUGH / PYREXIA | NO | NO | OXYGEN / ANTIBIOTICS | DISCHARGED |
| P06 | F | SOB / COUGH / PYREXIA | NO | NO | OXYGEN / ANTIBIOTICS / STEROIDS / REMDESEVIR | DISCHARGED |
| P07 | F | SOB / COUGH / PYREXIA | NO | No | OXYGEN / ANTIBIOTICS / STEROIDS / REMDESEVIR | DISCHARGED |
| P08 | F | COUGH / DIARRHEA / PYREXIA | YES | No | ANTIBIOTICS | DISCHARGED |
| P09 | M | DROWSY / PYREXIAL / VOMITING | ? | NO | NO DATA | DECEASED |
| P10 | F | FEVER / COUGH / SOB | NO | YES | OXYGEN / ANTIBIOTICS / STEROIDS / REMDESEVIR | DISCHARGED |
| P11 | F | PV BLEEDING | NO | NO | ANTIBIOTICS / STEROIDS | DISCHARGED |
| P12 | M | CP /<br>COUGH /<br>SOB | NO | No | OXYGEN /<br>ANTIBIOTICS /<br>STEROIDS /<br>REMDESEVIR | DISCHARGED |

**Table 2:**
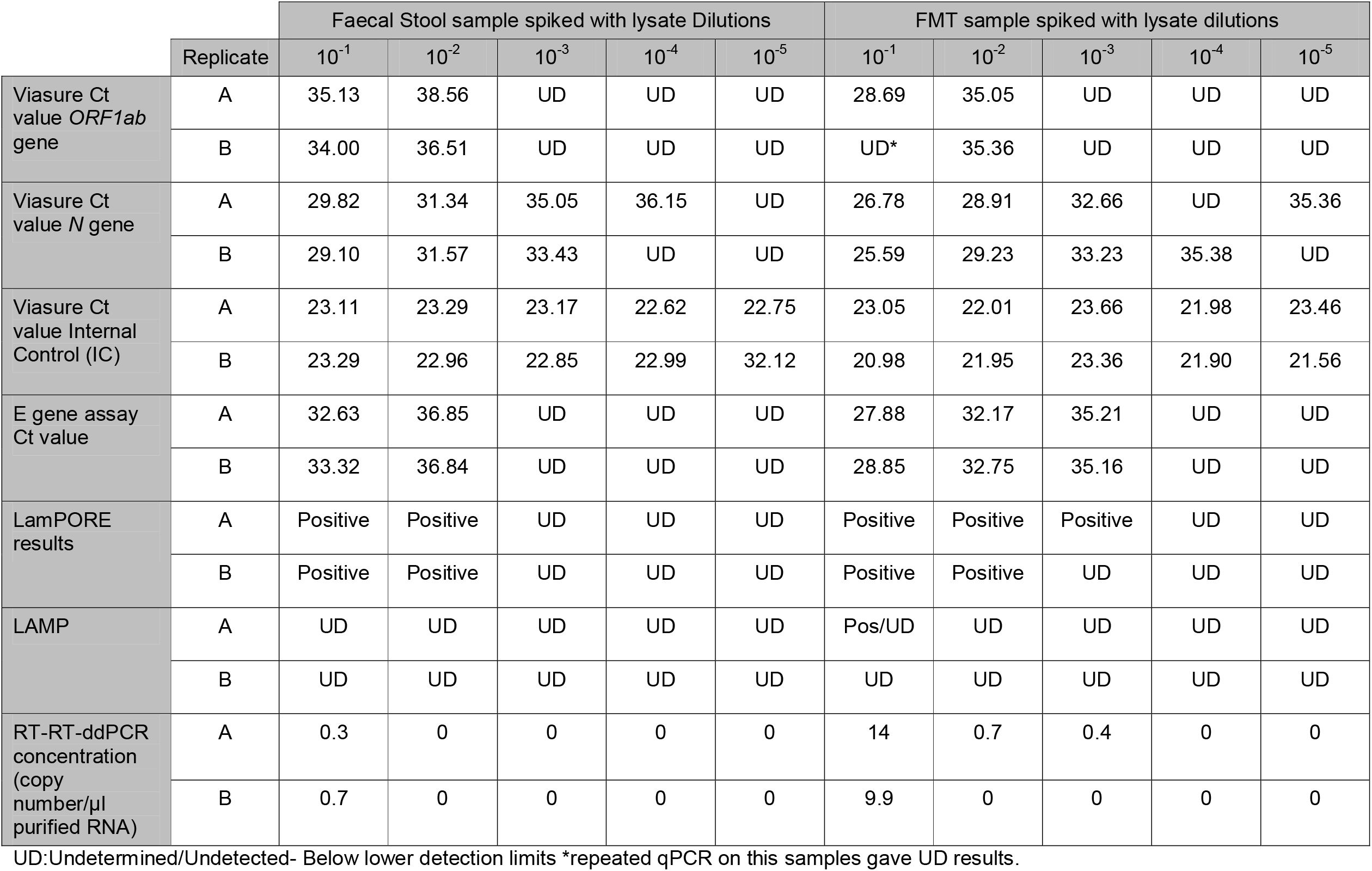
Amplification results of dilutions of Cell lysate spiked into Faecal and FMT samples.

Using the Viasure Real time PCR detection kit, amplification of the SARS-CoV-2 *orf1ab* and *N* genes were detected in the 10^−1^ and 10^−2^ lysate spiked faecal and FMT samples as well as the lysate extraction controls at all dilutions. Amplification in 10^−3^ to 10^−5^ concentrations for the lysate spiked samples was undetected. Similarly, using the WHO *E* gene Real-Time PCR detection assay, amplification of Envelope gene (*E* gene) of SARS-CoV-2.was detected in 10^−1^ and 10^−2^ lysate spike faecal samples and10^−1^ and 10^−3^ lysate spike FMT samples. Using the LamPORE SARS-CoV-2 protocol, amplification *orf1ab, E* and *N* genes were detected in 10^−1^ and 10^−2^ lysate spike faecal samples and10^−1^ and 10^−3^ lysate spike FMT samples. No signal was seen in any negative controls (Table 3).

**Table 3:** Amplification results of dilutions of RNA and Cell lysate extracted Bristol control samples

|  | Lysate extraction controls |  |  |  |  | RNA extraction controls |  |  |  |  |
| --- | --- | --- | --- | --- | --- | --- | --- | --- | --- | --- |
|  | 10<sup>-1</sup> | 10<sup>-2</sup> | 10<sup>-3</sup> | 10<sup>-4</sup> | 10<sup>-5</sup> | 10<sup>-1</sup> | 10<sup>-2</sup> | 10<sup>-3</sup> | 10<sup>-4</sup> | 10<sup>-5</sup> |
| Viasure<br>Ct value *ORF1ab*<br>gene | 19.93 | 26.14 | UD | UD | UD | UD | UD | UD | UD | UD |
| Viasure<br>Ct value *N* gene | 24.17 | 27.13 | 29.63 | 31.50 | 33.39 | 36.23* | UD | 31.29* | UD | UD |
| Viasure<br>Ct value Internal<br>Control (IC) | 22.43 | 22.10 | 21.20 | 21.11 | 22.72 | 22.97 | 22.40 | 21.38 | 22.27 | 22.19 |
| E gene assay Ct<br>value | 21.16 | 25.28 | 29.24 | 31.75 | 34.40 | UD | UD | UD | UD | UD |
| LamPORE<br>results | Positive | Positive | Positive | Positive | Positive | UD | UD | UD | UD | UD |
| Optigene LAMP<br>results | Positive | Positive | Positive | UD | UD | UD | UD | UD | UD | UD |
| RT-RT-ddPCR<br>concentration<br>(copy number/μl<br>purified RNA) | 1690 | 125 | 8.8 | 0.8 | 1 | 0 | 0 | 0 | 0 | 19 |

UD:Undetermined- Below lower detection limits
\*Inconclusive amplification

The results of the digital droplet RT-qPCR correlated negatively with N gene results from the ViaSure assay (Pearson’s moment correlation coefficient r= -0.89) indicating good correlation of Ct values with viral content as assessed by the dd-rt-PCR experiment. LAMP only detected 10-1 spiked lysate concentration within the stool samples, with no virus detected in any other samples, likely due to template degradation during RNA extraction.

### Patient derived samples

Of the 12 symptomatic COVID-19 patients who provided stool samples (Table 1, 4), 41.6% (5/12) had SARS CoV-2 RNA detectable in their stool via RT-QPCR, 5/12 (41.6%) via LamPORE and 6/12 (50%) via droplet digital qPCR. Of these patients 25% (3/12) reported GI symptoms and of note, none of the patients with RNA shedding in their stool reported GI symptoms. The average age of the 12 patients was 55 and the median age was also 55. The majority of the patients 92% (12/13) recovered from their illness and were discharged, with one patient dying from their infection.

**Table 4:** Hospitalised COVID-19 positive patient stool sample results using the Viasure Real-Time PCR SARS-CoV-2 detection kit.

| Patient Number | Viasure validated data | | | SARS-CoV-2 Detected | RT-ddPCR Concentration (copy number/μl purified RNA) |
| --- | --- | --- | --- | --- | --- |
|  | Ct value *ORF1ab* gene | Ct value *N* gene | Ct value IC |  |  |
| P01 | UD | UD | 24.69 | N | 0 |
| P02 | UD | UD | 23.23 | N | 0 |
| P03 | 28.02 | 24.33 | 24.59 | Y | 10.7 |
| P04 | UD | UD | 24.26 | N | 0 |
| P05 | 37.21 | 23.60 | 23.86 | Y | 0 |
| P06 | UD | UD | 23.89 | N | 0 |
| P07 | UD | UD | 23.81 | N | 0.8 |
| P08 | 32.75 | 23.24 | 24.04 | Y | 1 |
| P09 | 28.34 | 23.68 | 24.26 | Y | 8.9 |
| P10 | UD | UD | 24.04 | N | 0 |
| P11 | 34.10 | 23.09 | 23.43 | Y | 0.8 |
| P12 | UD | UD | 23.78 | N | 0 |
| Extraction Positive control | 20.98 | 19.91 | 23.56 | Y | 1278 |
| Extraction negative control | UD | UD | 23.43 | N | 0 |
| Viasure PCR positive control | 20.46 | 19.72 | 24.32 | Y | 295* |
| PCR negative control | UD | UD | 24.10 | N | 0 |

UD:Undetermined- Below lower detection limits
- *E* gene positive control 10<sup>-3</sup> ATCC control

We collected and analysed stool samples from a total of 97 ‘asymptomatic’ patients presenting to SWBH over a 3-month period. This included a mix of new presentations to the surgical admissions unit (SAU) under various surgical specialities and existing inpatients. The mean age of our cohort was 65 years (range 20 – 94), median of 68 years. This included 55 (57%) females and 42 (43%) males. The majority of our participants were White-British – accounting for 74% of the total, with smaller numbers of Asian-British and Black-British patients. The majority of our patients (74%) with various co-morbidities presented with abdominal pain and were admitted under general surgery. Smaller numbers were under the care of urology, trauma and orthopaedics and medicine. Approximately 22% (21) of our patients had a stoma (ileostomy or colostomy).

Eighty (80%) percent (78) of our cohort denied symptoms of COVID-19 in the two weeks leading to hospital attendance/admission. Of the remainder, 18 (19%) complained of suffering with diarrhoea in the last 14 days. None of our patients had a fever, new onset cough, anosmia or ageusia.

The majority of our patients had a nasopharyngeal swab taken on admission to the hospital as per local Trust protocol. 78 (80%) of these returned a negative swab result, 11 (11%) were rejected by the laboratory (inadequate labelling), and in 5 the swab was either not collected or declined by the patient. Only 1 patient had a positive swab result. Using QPCR, 0/96 nasopharyngeal swab negative patients had detectable SARS-CoV-2 virus in their stool samples. In one patient who was swab positive 25 days previously (27 days post symptom onset) there was detectable SARS CoV-2 RNA in his stool with Ct value of 32.37 (ORF1ab gene) and 23.23 (N gene).

## CONCLUSIONS

We have demonstrated with a CE-IVD marker two gene RT-qPCR assay, amplification was detected for both the *ORF1ab* and *N* genes in both faecal and FMT lysate spike replicates at 10^−1^ and 10^−2^ dilutions with lower Ct values detected with the more concentrated samples. Similar results were obtained using the E gene kit and reflected in the LamPORE and digital droplet experiments. We saw little amplification with LAMP testing, however as demonstrated via the RNA extraction results on a fragment analyser, RNA quality was poor and it is likely that passage through the extraction kit led to excessive template degradation. Commercial stool kits for LAMP testing in veterinary applications, suggesting further optimisation is needed for use on human stool samples.

The potential for faeco-oral asymptomatic transmission of SARS-CoV-2 has significant implications for FMT programmes globally. With FMT services on pause, treatment of patients with recurrent and refractory CDI remain sub-optimal with a likely consequential impact on morbidity, mortality and health care resource. Availability of a validated SARS-CoV-2 stool assay for donor screening would facilitate safe restart of FMT production and supply. Through our optimised methodology we have shown that using the VIASURE assay we can detect SARS-CoV-2 in stool samples containing more than 200 viral copies per gram. This is comparable to the widely used nasopharyngeal swab testing (14). As faecal viral shedding persists long after clearance from the upper respiratory tract (for up to 33 days in stool), the direct testing of faecal samples arguably is necessary. It is notable that in the spiking experiments whereas RNA was detectable in lysate spiked samples in neither donor stool nor prepared FMT was directly spiked RNA detectable. This is probably due to RNA destruction during the extraction process whereas in lysate the genetic material is protected within virions. Furthermore, for the first time, in the current work we have been able to demonstrate the efficacy of a Nanopore technology platform in faecal samples. This exciting development opens the possibility of rapid near-subject testing of stool samples with potential for applications beyond the current COVID-19 pandemic.

Our limited patient data demonstrates that about 40% of symptomatic patients admitted with acute COVID-19 test positive for SARS-CoV-2 in faecal samples and this is congruent with other published data (6). Although the main impetus behind our work is to ensure that we have a suitable stool assay to resume our national FMT service, the finding of SARS-CoV-2 in stool highlights the intriguing potential role of the gastro-intestinal tract in the pathogenesis of the disease. We also have shown that SARS-CoV-2 cannot be detected in the stool of asymptomatic patients presenting with acute surgical emergencies in an area with high incidence of SARS-CoV-2. While true faecal-oral transmission and infectivity of stool samples from COVID-19 patients has yet to be confirmed, there may be sufficient circumstantial evidence to suggest that this is likely to be the case in certain circumstances.

In previous work examining antibody development against SARS-CoV2 about a quarter of hospital staff had diarrhoea and there was an association between this and the likelihood of antibody detection in serum (18). There is accumulating evidence that the gut mounts an active immunoglobulin response to the virus (19) and it is suggested that the gut IgA response may impact on the efficacy of the body’s response to vaccination. In vitro work from several laboratories have shown that intestinal epithelium is readily infected suggesting that the intestine is a potential site of SARS-CoV-2 replication (20) with upregulation of viral response genes. Moreover, there is increasing evidence that SARS-CoV2 infection is associated with intestinal inflammation based on measurement of faecal calprotectin(21). In contrast, a large retrospective cohort reported an intriguing suggestion that gastro-intestinal involvement may be associated with a more benign outcome of infection (22). Although these data are subject to question in light of the retrospective study design there was a signal of a comparatively ‘anti-inflammatory’ peripheral cytokine response in patients with gastrointestinal symptoms.

Interestingly, in our study of asymptomatic patients presenting to an acute surgical unit during the peak of the 2^nd^ wave of SARS-CoV-2 infection in the UK, we found no positivity either via NP swab or in stool, despite a proportion of them having diarrhoea on admission. A single patient was identified who was previously positive for SARS-CoV-2 who had detectable virus in their stool 25 days after initial positivity. Together these findings suggest that faecal transmission in the absence of a positive nasopharyngeal swab is unlikely to be an issue, and that faecal transmission is not an important route in SARS-CoV-2 in asymptomatic patients.

The practice of FMT, while revolutionary for the treatment of CDI (9) and showing promise in IBD (23), has recently been stalled as a result of transmission of infective pathogens to patients who came to harm as a result(10). Recent cases include drug resistant *Escherichia coli* (Enteropathogenic *Escherichia coli* and *Shigatoxin-producing Escherichia coli*) and related to inadequate screening of donated stool samples.

It is therefore imperative that donated FMT stool samples are carefully screened for SARS-CoV-2 and only used once tested negative (12). The Chinese University of Hong Kong has an active FMT program and, similar to our approach, they have adopted a similar approach of careful enhanced donor screening and PCR testing of stool samples in order to perform FMT safely (24).

## Data Availability

All data is available on reasonable request to the authors

## Acknowledgments

None

